# The Prevalence of Self-Medication and Its Associated Factors Among the Residents of Kandahar, Afghanistan

**DOI:** 10.64898/2026.02.05.26345701

**Authors:** Atta Ur Rahman Arian, Mohibullah Mako, Nasib Ahmad Shaikhal, Najeebullah Rafiqi, Bilal Ahmad Rahimi

## Abstract

**Background:** The World Health Organization (WHO) defines self-medication as taking medicine by someone to treat a health problem on their own, without a professional or doctor consultation. Self-medication is a growing public health concern globally, including in Afghanistan. This study aims to assess the prevalence of self-medication and its associated factors among the residents of Kandahar city.

**Methodology:** An analytical cross-sectional study was conducted in 12 districts of Kandahar city. A total of 401 adults were randomly selected and interviewed at private pharmacies using a structured questionnaire. Data collection was done from June 22 to July 17, 2025. The data was analyzed using SPSS version 26, with descriptive and analytical analyses, considering a *p*-value below 0.05 as statistically significant.

**Results:** A high prevalence (71.1%) of self-medication was observed among the residents of Kandahar. More than one type of medication was used by 47.1% participants, with paracetamol being the second most commonly used (18%). More than half (54.1%) of medicines were used for multiple symptoms. The specific reasons for self-medication were high doctor fees (18.5%) and previous experience with similar symptoms (13.7%). Pharmacies were identified as the primary source of self-medication (69.8%). Statistically significant factors associated with self-medication were found to be lack of awareness about self-medication (*p*-value = 0.003), easy access to pharmacies (*p*-value = <0.001), and the presence of symptoms (*p*-value = <0.001).

**Conclusion:** Self-medication was highly prevalent in Kandahar, with key associated factors such as lack of awareness, easy access to pharmacies, and the presence of symptoms. It is recommended to implement health education programs to increase awareness about the adverse effects of self-medication. Strict implication of rules and regulations is required to control the sales of medicines without prescription in pharmacies.

## Introduction

According to the World Health Organization (WHO), self-medication refers to a situation in which an individual relies on themselves for diagnosis, prescription, or treatment without a professional medical instruction [1]. Self-medication has become a common practice in many communities, where people choose to treat their health issues on their own, without healthcare professional consultation. Self-medication is considered a serious problem worldwide [2]. Recent research has shown that nearly half of the world population relies on self-medication, with an overall global prevalence of 48.6%. Based on continent-level analysis, the highest prevalence was found in Asia (53%), followed by America (47.8%), Africa (41.5%), and Europe (40.8%). The highest prevalence of self-medication was reported among students (54.5%), and the lowest (32.5%) was linked to healthcare workers [3]. Similarly, self-medication is at its peak in the neighboring countries of Afghanistan. Studies have reported the highest prevalence of self-medication among individuals in Pakistan (84%) [4], India (64.4%) [5], China (58.60%) [6], and Iran (53%) [7]. Self-medication is an increasing public health concern. It is influenced by a variety of associated factors, including an individual’s education level, parents’ educational status, place of residence (living in an urban area), easy access to pharmacies, and pressure from peers or family [8]. Besides the education, gender and high income are also linked with self-medication [9]. In addition to that, people often chose self-medication because they had experienced similar health conditions and felt sure of managing them, and the urgency of their symptoms pushed them to self-medication [10]. A study conducted in Nangarhar province of Afghanistan highlighted a 62.6% prevalence of self-medication, with key associated factors such as easy availability of obtaining medicine, high healthcare costs, experience of similar symptoms before, low awareness about the negative effects of drug usage, and not being serious about the symptoms [11]. Self-medication with antibiotics is considered high in Kabul, with a prevalence of 59.1%, and earlier experiences of the same diseases, lack of money to see a doctor, were the primary reasons for self-medication [12]. Other than this, nearly 34.9% of the patients were relying on self-medication in a clinic in Kabul, and lack of time, previous experiences, and saving money were defined as associated factors for self-medication [13]. Another study among university students shows that 25.16% of the students practiced self-medication. From those, 59.2% of the medicines were found that should only be taken with a doctor’s prescription [14].

Collectively, all over the world, including Afghanistan, numerous studies have identified the prevalence of self-medication and its associated factors. Since no or very limited studies have been conducted to assess the prevalence of self-medication and its associated factors in Kandahar province, therefore, the researchers considered this as a gap to study this issue. The findings will benefit public health authorities, health-care providers, policy makers, community pharmacists, researchers, and the general population. The results can inform policies, educational programs, and strategies in promoting the safe and responsible use of medications.

## Methodology

### Study Design

This study employed an analytical cross-sectional research design to assess the prevalence of self-medication and associated factors among the residents of Kandahar city. This design was considered appropriate because it allows for gathering standardized data from a large group of participants at one specific time.

### Study site

Kandahar is located in the southwestern part of Afghanistan and is the second-largest city in the country. This study was conducted across 12 districts within Kandahar City. These districts were selected randomly to ensure representation from different geographic, demographic, and socioeconomic areas within the city. Private pharmacies were selected as research sites because they were the right sources where people obtain medicine for self-medication.

### Study Population and Sampling

The study population included people aged above 18 years and residing in Kandahar. Participants were also eligible if they had obtained medicine without a doctor’s prescription in the past three months. A total of 401 participants were randomly selected from the population of 12 districts for data collection. A random sampling technique was employed to ensure that every individual had an equal chance to participate, which helps to produce a representative sample of the population.

### Date Collection

Data were collected using a self-structured questionnaire developed after reviewing the relevant literature on self-medication. The questionnaire consisted of five parts, including Demographic information, experiences with obtaining and using medicines without a prescription, reasons behind self-medication, awareness about the risks linked to self-medication,

The data were gathered through face-to-face interviews conducted at private pharmacies across 12 selected districts of Kandahar City. Participants were individuals who were obtaining medicines at private pharmacies at the time of data collection. Before each interview, participants were informed about the purpose of the study, and their written consent was obtained. Moreover, Data collection took place during the period from June 22 to July 17, 2025, to reach the required sample size of 401 participants.

### Data Analysis

The collected data were analyzed using SPSS software version 26. Descriptive and analytical statistics were used to show the prevalence and also assess factors associated with non-prescription drug use. Descriptive statistics were used to describe the distribution of each variable using frequencies and percentages. Chi-square test was performed to assess the association between independent variables and self-medication. A *p-*value <0.05 was considered a significant association between variables.

### Ethical Consideration

This research was conducted adhering to ethical considerations. Before taking part in the study, participants were clearly informed about the purpose of the study, and their consent was obtained. They were assured that their responses would remain confidential and would be used only for research purposes. In order to begin the research, an Institutional Review Board (IRB) approval of Kandahar University was obtained. Also, all ethical principles of declaration of Helsinki were strictly followed throughout the data collection and reporting process.

## Results

This study aimed to assess the prevalence of self-medication and its associated factors among the residents of Kandahar. To achieve the initial understanding of the data, a descriptive analysis was conducted to describe the frequency and percentage of the key variables related to self-medication practices. The demographic information of the participants, age, such as gender, education level, and occupation, is presented in Table 1. The analysis shows that majority (93.0%) of the respondents were male and only (7%) were female. A large number (41.6%) of the study participants were illiterate, indicating low educational achievement in the sample, with 27.7% having primary education and only 13% having bachelor degree. Most of the participants (74.3%) worked in the private sector, while 25.7% were government employees. This indicates that the majority of the participants were male, had low educational attainment, and were private sector employees.

**Table 1.** Demographic data of the study participants.

| Variable | Category | Frequency (n=401) | Percentage (%) |
| --- | --- | --- | --- |
| Age (in years) | 18–40 | 287 | 71.6 |
|  | 40–60 | 91 | 22.7 |
|  | >60 | 23 | 5.7 |
| Gender | Male | 371 | 92.5 |
|  | Female | 30 | 7.5 |
| Education level | Illiterate | 167 | 41.6 |
|  | Literate | 234 | 58.4 |
| Occupation | Government employee | 103 | 25.7 |
|  | Private sector employee | 298 | 74.3 |

Table 2 shows the practices of self-medication among the respondents. The results indicate that 71% of the respondents reported practicing self-medication, while 29% were using medicine with a doctor’s prescription. Over the past three months, 12% had purchased medicine once, 35.2% occasionally, and 23.9% frequently, while 28.9% had not purchased any medicine. Regarding the types of medications used without prescription, nearly half (47.1%) of the study participants had used more than one type of medicines, with paracetamol used by 18% and antibiotics by 4.2% of study participants; among them, 28.9% had not practiced self-medication. The symptoms for which respondents took medicine were mostly multiple complaints (54.1%), followed by headache (5.5%) and stomach problems (5.4%). Collectively, a large portion of the population was engaged in using medicine without professional guidance. The purchase of medication was high, more than one type of medicine were used by the participants without a prescription, and self-medication was experienced for more than one symptom.

**Table 2.**
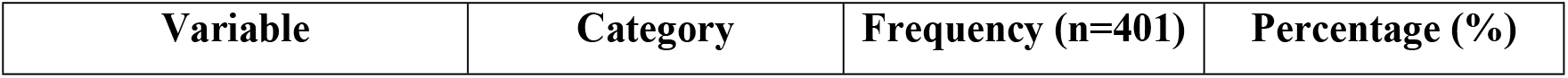
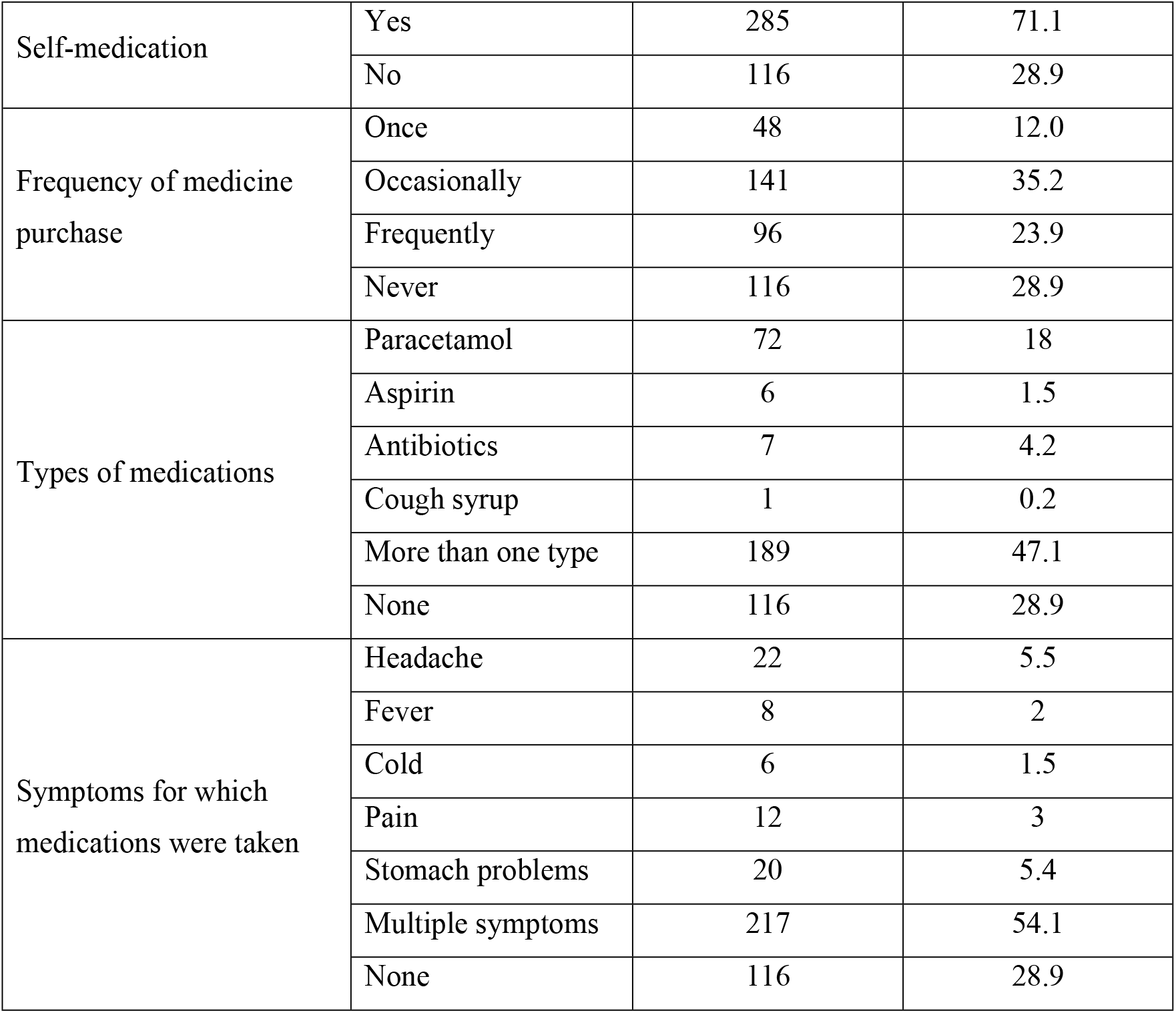
Self-medication practices among the study participants.

Table 3 shows the reasons and access to non-prescription medication use. This table reveals that self-medication is influenced by various factors. The most common reported specific reason for self-medication was high fees of the doctors (18.5%), suggesting that economic of financial situations play a major role in self-medication practices. This was followed by previous experience with similar illnesses (13.7%) and not being serious about the illness 8.7%. Saving time (5.5%) and advice from pharmacists (4.2%) were less commonly reported, and 20.4% of the study participants reported having more than one reason. The findings also show that most (69.8%) of the study participants obtained medications from pharmacies, highlighting easy access to medicines, while very few relied on friends, family, and the internet.

**Table 3.** Reasons and access to self-medication among the study participants.

| Variable | Category | Frequency<br>(n=401) | Percentage (%) |
| --- | --- | --- | --- |
| Reasons for self-medication | Saving time | 22 | 5.5 |
|  | High doctor's fee | 74 | 18.5 |
|  | Minor illness | 35 | 8.7 |
|  | Previous experience | 55 | 13.7 |
|  | Advice from a pharmacist | 17 | 4.2 |
|  | More than one reason | 82 | 20.4 |
|  | None | 116 | 28.9 |
| Access to self-medication | Pharmacies | 280 | 69.8 |
|  | Friends | 2 | 0.5 |
|  | Family members | 2 | 0.5 |
|  | Internet | 1 | 0.2 |
|  | None | 116 | 28.9 |

**Note:** Due to multiple responses, percentages may not equal to 100%.

Table 4 reveals the experience, awareness, and opinion about self-medication among the respondents. The findings indicate that 33.9% of the participants experienced problems due to self-medication, while the majority (66.1%) did not. Over half (52.6%) of the respondents were aware of the harmful effects of drug usage without prescription. When asked about the safety of self-medication, the majority (88.5%) perceived it as unsafe, whereas a very small proportion considered it safe. This suggests that self-medication is prevalent, and most participants understand its potential risks.

**Table 4.**
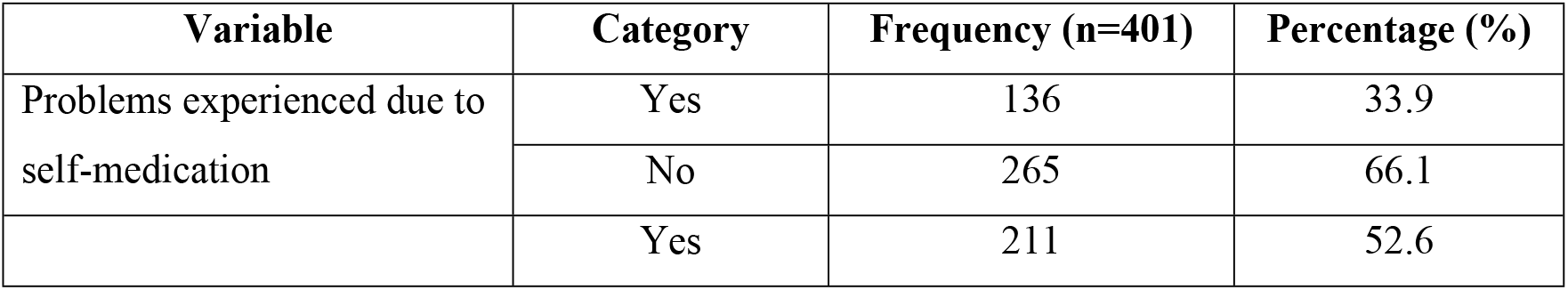
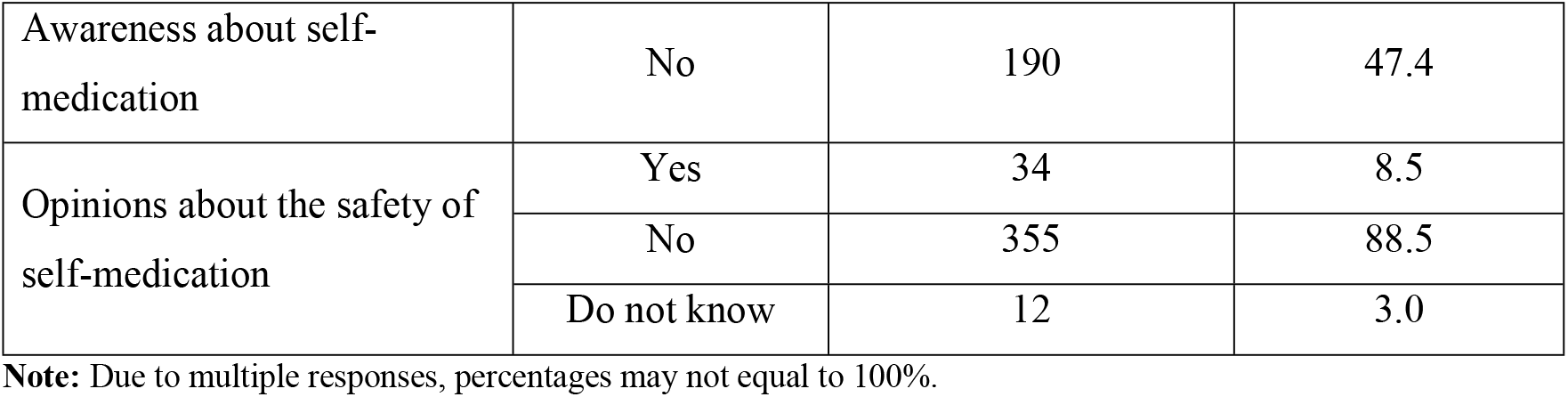
Awareness, experience, and perception regarding self-medication among the study participants.

Table 5 presents the results of Chi-square test to assess the factors associated with self-medication among the study participants. A statistically significant association was found between awareness about self-medication and using a drug without a prescription (*p*-value = 0.003). Participants with lower awareness demonstrated a significantly higher prevalence of self-medication compared to those with adequate awareness, indicating that a lack of awareness is an important factor contributing to self-medication. Easy access to pharmacies was found to be significantly associated with non-prescription drug use (*p*-value = <0.001). The majority of participants obtained medicines directly from pharmacies, highlighting the notable role of pharmacies in facilitating self-medication without prescription. Finally, a highly significant association was observed between self-medication and the presence of symptoms (*p*-value = <0.001). Almost every respondent was experiencing symptoms when obtaining medication without a doctor’s instruction. Overall, the data clearly suggests that self-medication is highly influenced by lack of awareness, easy access to pharmacies, and the presence of symptoms or signs of illness.

**Table 5.** Chi-square test to show statistically significant factors associated with self-medication among the study participants.

| **Variables** | **Self-medicaition**<br>(n=285), n<br>(%) | **No self-medication**<br>(n=116), n<br>(%) | **Total**<br>(n=401), n<br>(%) | ***P-value*** |
| --- | --- | --- | --- | --- |
| Awareness about self-medication |  |  |  |  |
| Yes | 138 (65.4) | 73 (34.6) | 211 (100) | 0.003 |
| No | 147 (77.4) | 43 (22.6) | 190 (100) |  |
| Access to self-medication |  |  |  |  |
| Pharmacy | 273 (97.5) | 7 (2.5) | 280 (100) | <0.001 |
| Outside pharmacy | 12 (9.9) | 109 (90.1) | 121 (100) |  |
| Presence of symptoms |  |  |  |  |
| Having symptoms | 284 (99.6) | 1 (0.4) | 285 (100) | <0.001 |
| No symptoms | 1 (0.9) | 115 (99.1) | 116 |  |

## Discussion

This study was intended to assess the prevalence of self-medication and its associated factors among the residents of Kandahar. The results revealed a notably high level of medication use without a proper doctor’s prescription. The majority of the participants reported practicing medication without a prescription, with a prevalence of 71.1%. Similarly, a recent systematic review reported an overall self-medication prevalence of 70.1% among university students [15], [16]. In Iran, the practice of non-prescription drug use rate was reported 89.6%, which is higher than the prevalence of this study [17]. The observed prevalence was higher in several studies conducted in India (66.4%) [18], Myanmar (57.8%) [19], and Jordan (56.9%) [20]. The prevalence of self-medication was also higher from the studies previously conducted in this context, Afghanistan [11], [12], [13], [14].

The current study identified that 47.1% of participants were using more than one type of medication. Similarly, in Pakistan, multiple kinds of medicine were obtained from the community pharmacies [21]. Paracetamol was the second most commonly used medication among the participants, with a prevalence of 18%, which is lower than the prevalence reported in India (52%) [22]. In the present study majority of participants 54.1% reported obtaining medication for multiple symptoms. Comparable to findings reported in other studies, where self-medication was practiced for more than one symptom, such as headache, fever, pain, cold, stress, and menstrual problems [23], [24]. In addition to these factors, another study conducted in Myanmar identified other reasons such as cough, ache in throat, teeth, bones, muscles, stomach symptoms, and high blood pressure [19].

A study conducted in Nangahar, high healthcare costs 57.2%, and earlier experiences with similar symptoms 49.8% were major reasons behind self-medication [11]. Differ from the findings of the current study, where high health-care fees 18.5% and previous experiences of the problem 13.7% was found to be specific reported reasons behind self-medication. In the present study, 69.8% of medicines used for self-treatment were taken directly from pharmacies. This proportion is lower than reported in a student-based study, in which 91.7% of students obtained medicines from pharmacies [25].

The majority (66.9%) of participants stated not experiencing problems after practicing non-prescription medicine, which is lower than a study conducted in Bangladesh, where approximately 93.08 % of participants did not report any side effects after using antibiotics without a prescription [26]. While 33.1% reported not being negatively affected after self-medication. This prevalence is a bit higher than an elderly-focused study (26.7%), but significantly lower than another study that reported a 75% prevalence of the side effects of self-medication [27]. More than half of the participants (52.6%) were aware of the adverse effects of self-medication. However, this awareness level is lower than the studies conducted in Nigeria (82.3%) [28] and Italy (66.8%) [29]. In contrast, another study reported that only 45.6% of participants were aware of the harmful effects of non-prescription drug use, which is lower than the awareness level found in the present study [30]. Moreover, 88.5 % of the respondents perceived self-medication as unsafe; this proportion is higher than previously reported in Saudi Arabia (65.9%) [31]

The current study investigated self-medication practices and their associated factors among the participants. The results highlighted that low awareness regarding self-medication was significantly associated with non-prescription drug use. This finding aligns with research conducted in Nangarhar province of Afghanistan, where a lack of awareness was linked to self-medication [11]. Similarly, a study from Indian Punjab reported that 11.5% of participants had low awareness about self-medication [32]. These results are also consistent with previous research showing a significant relationship between health literacy and self-medication [33], [34].

Easy access to medication was significantly associated with nonprescription drug use in the current study. Within this variable, pharmacies were identified as the major source. Nearly all participants who practiced self-medication obtained medicines directly from pharmacies, indicating the central role of pharmacies in facilitating drugs without prescription. These findings are consistent with previous research conducted in similar contexts. For example, a study from Afghanistan reported that pharmacies were the most common source of self-medication, with 85.9% of participants obtaining antibiotics from pharmacies for self-treatment [12]. Moreover, a large survey conducted in Nangarhar province found that easy access to pharmacies 64.7% was one of the main reasons for self-medication among adults [11]. Similar patterns have also been reported in neighboring countries, including Iran [35] and India [36]. Additionally, in Alexandria, Egypt, nearly 81% of medicines used for self-treatment were directly purchased from pharmacies without a prescription [37].

Finally, the presence of symptoms were statistically significant factor associated with self-medication. This consist with previous research in Afghanistan, where runny nose, sore throat, and cold were the primary symptoms leading to self-medication [12]. In another study from Afghanistan, pain relief, headache, fever, and stomach problems were identified as the common reasons [11]. The findings also align with previous research across several countries. Among Iranian students, headache, body pain, and cold or cough were the most frequently self-treated symptoms [38]. In Pakistan, fever, cold, and diarrhea were reported as the main symptoms leading to self-medication [39]. Similarly, in Bangladesh, cold, headache, and stomach problems were commonly self-treated by the participants [40]. In addition to cold and stomach problems, heart disease was also identified as a symptom for which self-medication was frequently used in China [41].

In summary, a high prevalence of self-medication was observed in Kandahar, with associated factors such as lack of awareness about non-prescription drug use, easy access to pharmacies, and the presence of symptoms. These factors were also observed in a previous study conducted in Nangahar, indicating that self-medication is a significant public health concern across different regions of Afghanistan. It highlights the need for public health programs aimed at increasing awareness about the risks of self-medication, and rules to be developed to control the sales of non-prescription medicines in pharmacies.

## Limitations

There were limitations in our study. First, the cross-sectional design of the study. It can show associations between factors and self-medication, but cannot prove causality. Second, this study only assessed the prevalence of self-medication and its associated factors among urban residents. Future research could provide a better understanding by exploring self-medication practices in rural areas as well. Third, predominance of male participants. Collecting data from the females would ensure more accurate and generalizable results.

## Conclusion and recommendations

This study found a high prevalence of self-medication among residents of Kandahar. Self-medication was significantly associated with low awareness about self-medication, easy access to medicines through pharmacies, and the presence of symptoms. The findings suggest that self-medication is a significant public health concern in Kandahar. It is recommended that public awareness programs about the harmful effects of self-medication be employed, and rules and regulations are required to be strictly followed to control the sale of non-prescription medicines. Based on the findings, the health education program implementation is recommended to increase awareness about the risks of self-medication. There is a need for stronger application of regulations to control the sale of medicines without a prescription. Regular monitoring of pharmacies is recommended to reduce inappropriate access to non-prescribed drugs. Also, pharmacists are advised to provide proper guidance to customers and refer them to healthcare facilities when symptoms require medical attention.

## Acknowledgement

The authors would like to express gratitude to all the participants who took part in this study. Appreciation also goes to local health centers and pharmacies for their cooperation during data collection. Finally, a special thanks to Mr. Hilal Ahmad Tareen, whose kind assistance in data collection played a crucial role in the completion of this research.

## AUTHOR CONTRIBUTIONS

- AURA: Conceptualization, Investigation, Formal Analysis, Writing – Original Draft, Project Administration.
- MM: Methodology, Supervision, Writing – Review & Editing.
- NAS: Validation, Supervision, Writing – Review & Editing.
- NR: Supervision, Formal analysis, Writing – Review & Editing.
- BAR: Supervision, Methodology, Formal analysis, Writing – Review & Editing, Resources.

## Funding

The authors received no specific funding for this work.

## Conflict of Interest

The authors have declared that no competing interests exist.

## Data Availability Statement

All relevant data are within the manuscript and its Supporting Information files.

## Ethical Statement

This study was approved by the Kandahar University Institutional Review Board (Reference No: KDRU-IRB-016. Dated 5/23/2025).

## Informed Consent

Written informed consent was obtained from all participants after a detailed explanation of study objectives, procedures, voluntary participation, withdrawal rights, confidentiality, and data use.

## Notes

### Competing Interest Statement

The authors have declared no competing interest.

### Funding Statement

The author(s) received no specific funding for this work.

